# Life Expectancy in the United States Returns to PrePandemic Levels – an Update

**DOI:** 10.1101/2025.05.29.25327935

**Authors:** Harry Wetzler

## Abstract

From 2019 to 2021, U.S. life expectancy at birth fell by roughly 2.45 years to the lowest level since 1996. Although life expectancy rose in 2022 and 2023, it did not fully return to its 2019 level. Now that 2024 death reports have stabilized, I constructed abridged life tables for 2019– 2024 using CDC WONDER Multiple Cause of Death mortality data and Census Bureau Vintage population estimates to assess recent trends. This is an update of a report posted on June 2, 2025.

In 2024, life expectancy at birth climbed by nearly 0.6 years over 2023 to 79.12 years, while life expectancy at age 65 increased by approximately 0.2 years to 19.87 years. These 2024 gains have almost offset the declines experienced during the COVID-19 pandemic. However, US mortality continues to exceed that in other high-income countries.

## Introduction

US life expectancies at birth declined by 2.1 years for females and 2.8 years for males between 2019 and 2021.^1,2^ The resultant life expectancies in 2021 were the lowest seen since 1996.^3^ Life expectancies at age 65 also declined, 1.1 years for females and 1.2 years for males between 2019 and 2021. My previous results and those from CDC/NCHS revealed that approximately 76% of the 2019-2021 declines were regained in 2022 and 2023.^4,5,6,7^ The purpose of this study was to estimate US life expectancies for 2024. This study is an update of a report posted on June 2, 2025.^8^ Reported 2024 death totals have been stable since July 19, 2025; this revision is based on totals reported through September 20, 2025.

## Methods

Death data for 2019-2024 by year, gender, and 5-year age groups were obtained from the US Centers for Disease Control and Prevention (CDC) Wide-ranging ONline Data for Epidemiologic Research (WONDER) Multiple Cause of Death database that is updated weekly.^9^ This report includes deaths occurring through September 20, 2025. Because data were anonymized and publicly available, approval by an institutional review board and informed consent were not required in accordance with 45 CFR §46.

Two notable changes occurred following the previous report. First, weekly US death totals have not changed since August 16, 2025. Second, on September 30, 2025, the U.S. Census Bureau released the 2010-2020 Intercensal Estimates.^10^ These revise the prior decade’s estimates to align with the most current census and are considered the preferred series of data for that decade.

Population data for July 1, 2020-2024 came from the Vintage 2024 estimates.^11^ An abridged life table template from Public Health England (PHE) was used for life expectancy and standard error calculations.^12^ The PHE template uses 0.120 for the fraction of life lived by decedents dying in their first year. This was changed to 0.126, the average of the values used in the 2019-2021 US life tables. Another nuance of the PHE template is the method for obtaining the years lived in the final open ended age interval for ages 90+. This is calculated by dividing the number surviving by the death rate in the interval. The CDC/NCHS life tables use a blend of vital statistics reported deaths and Medicare data for the oldest ages because Medicare enrollees must have proof of age to enroll and are considered to be more accurate.^7^ Every year the age at death is not stated for a small number of deaths – 19 females and 38 males in 2024. These deaths were allocated to the <1 year age group to provide conservative estimates.

Life expectancy estimates obtained from CDC WONDER death and U.S. Census Bureau population data for 2019-2024 were compared to published values from the Centers for Disease Control (CDC) National Center for Health Statistics (NCHS).

Chiang’s formula was used to calculate the standard error of life expectancy.^13^ I followed the Strengthening the Reporting of Observational Studies in Epidemiology (STROBE) reporting guidelines.

## Results

Through September 20, 2025, 3,072,666 deaths were reported in the US for 2024, a 0.59% decrease from 2023. The changes for females and males were -0.04% and -1.09% respectively. The August 16, 2025 death total is 882 more than the projected total in the previous version of this report.

Table 1 lists life expectancies at birth for the published US life tables (CDC/NCHS) and those calculated in this study.

**Table 1.** US Life Expectancies at Birth.

|  |  | Year |  |  |  |  |  |
| --- | --- | --- | --- | --- | --- | --- | --- |
|  |  | 2019 | 2020 | 2021 | 2022 | 2023 | 2024 |
| Total | CDC/NCHS | 78.85 | 76.99 | 76.37 | 77.46 | 78.42 | -- |
|  | This study | 79.12 | 77.01 | 76.41 | 77.52 | 78.53 | 79.12 |
| Female | CDC/NCHS | 81.39 | 79.88 | 79.33 | 80.20 | 81.08 | -- |
|  | This study | 81.66 | 79.81 | 79.36 | 80.24 | 81.16 | 81.57 |
| Male | CDC/NCHS | 76.31 | 74.19 | 73.55 | 74.80 | 75.82 | -- |
|  | This study | 76.56 | 74.33 | 73.59 | 74.87 | 75.95 | 76.71 |

The 2019 estimates above are 0.10, 0.12, and 0.10 years less for the Total, Female, and Male categories compared to the previous estimates. ^8^The 2024 estimates for the Total and Male categories are each 0.01 year lower and the Female results are the same as the previous estimates. With one exception, the results in this study consistently exceed the CDC/NCHS values. The largest discrepancies occurred in the year 2019 estimates. The median difference for years 2020-2023 was 0.05 years.

Table 2 lists life expectancies at age 65 for the published US life tables and those calculated in this study.

**Table 2.** US Life Expectancies at Age 65.

|  |  | Year |  |  |  |  |  |
| --- | --- | --- | --- | --- | --- | --- | --- |
|  |  | 2019 | 2020 | 2021 | 2022 | 2023 | 2024 |
| Total | CDC/NCHS | 19.59 | 18.46 | 18.42 | 18.91 | 19.52 | -- |
|  | This study | 19.92 | 18.42 | 18.51 | 19.00 | 19.66 | 19.87 |
| Female | CDC/NCHS | 20.82 | 19.78 | 19.74 | 20.17 | 20.71 | -- |
|  | This study | 21.12 | 19.68 | 19.81 | 20.20 | 20.81 | 20.99 |
| Male | CDC/NCHS | 18.20 | 17.03 | 16.99 | 17.54 | 18.19 | -- |
|  | This study | 18.50 | 17.06 | 17.09 | 17.67 | 18.37 | 18.62 |

The 2019 estimates above are 0.13, 0.17, and 0.15 years less for the Total, Female, and Male categories compared to the previous estimates but those for 2024 are unchanged.^8^ The results in this study again generally exceed the CDC/NCHS values. The median difference for years 2020-2023 was 0.085 years. The Chiang standard errors for the 2024 estimates for the total population are 0.008 years at birth and 0.006 years at age 65. For females the standard errors are 0.011 years at birth and 0.008 years at age 65 and for males, 0.012 and 0.008.

## Discussion

Life expectancies in the US continued to increase in 2024 after starting to rise in 2022. With the gains made through 2024, the pandemic era declines have been erased for males and nearly erased for females. Despite these recent increases in life expectancy and the concomitant reductions in mortality, it is noteworthy that an estimated 1,366,642 excess deaths occurred in the US between February 1, 2020 and December 31, 2022.^14^ Furthermore, Bor and colleagues recently estimated that more than 3.6 million excess deaths occurred in the US in 2020-2023 compared to 21 other high-income countries.^15^

Comparing this study’s results with the “official” CDC/NCHS results for years 2019-2023 provides a gauge of this study’s accuracy. For 2020-2023, the life expectancy differences at birth were all 0.15 years or less. The greatest difference in life expectancy at age 65 was 0.18 years. The 2019 differences are greater largely due to the methods used for determining life expectancy in the last interval. Specifically, in 2019 the life expectancies at age 90 in this study exceeded those in the US life tables by an average of 20.3% while the average for 2020 and 2021 was 2.1%. Since the same method was used for each year in this study, it is unclear why the results for 2019 are so different.

Hanley raised the concern that reporting life expectancy to two decimal places implies nonexistent precision.^16^ He proposed a simple formula for estimating the standard error of life expectancy at birth. Comparing standard errors computed using Chiang’s method with those based on Hanley’s approach, the difference never exceeds 0.002 years. This close agreement suggests that observed increases in life expectancy between 2023 and 2024 reflect genuine gains.

Recent increases in US life expectancy may not be sustained. The One Big Beautiful Bill Act (P.L. 119-21) signed into law on July 4, 2025 may increase mortality in the US. The University of Pennsylvania and Yale University suggest that over 51,000 preventable deaths will occur each year due to the law’s provisions.^17^ In addition, cuts in the Supplemental Nutrition Assistance Program could add about 6,000 deaths annually.^18^ Adding 57,000 deaths with the existing age distribution of deaths would reduce US life expectancy by 0.2 years.

## Conclusion

US life expectancies continued to increase in 2024, and pre-pandemic levels have been largely achieved. It remains to be seen whether the 2019 level can be surpassed in the future. US mortality continues to exceed that in other high-income countries.

## Data Availability

All data produced in the present study are available upon reasonable request to the author.

https://wonder.cdc.gov/mcd-icd10-provisional.html

https://www.census.gov/newsroom/press-kits/2020/population-estimates-detailed.html

https://www.census.gov/data/tables/time-series/demo/popest/2020s-national-detail.html

https://www.census.gov/data/tables/time-series/demo/popest/intercensal-2010-2020-national-detail.html

